# Correlations between kidney and heart function bioindicators and the expressions of Toll-Like, ACE2, and NRP-1 receptors in COVID-19

**DOI:** 10.1101/2022.04.08.22273322

**Authors:** Rabab Hussain Sultan, Maged Abdallah, Tarek Mohamed Ali, Hebatallah Hany Assal, Amr E. Ahmed, Basem H Elesawy, Osama M. Ahmed

**Author notes:** Corresponding author: Rabab Hussain Sultan, 00201029819222.

## Abstract

**Background:** COVID-19 impacts the cardiovascular system resulting in myocardial damage and also affects the kidneys leading to renal dysfunction. This effect is mostly through the binding with angiotensin-converting enzyme-2 (ACE2) and Neuropilin-1(NRP-l) receptors. Toll-Like Receptors (TLRs) typically combine with microbial pathogens and provoke an inflammatory response.

**Aim:** This work aims to compare the changes in kidney and heart function bioindicators and expressions of TLRs (TLR2 and TLR2) as well as ACE2 and NRP-l receptors in moderate and severe COVID-19 patients. The correlations between kidney and heart function bioindicators and expressions of these receptors are also studied.

**Patients and Methods:** In this study, 50 healthy control and 100 COVID-19 patients (55 male and 45 female) were enrolled. According to WHO guidelines, these participants were divided into severe (50 cases) and moderate (50 cases). Serum creatinine, blood urea, CKMB, LDH, and Troponin I were estimated. We measured the gene expression for Toll-Like Receptors (TLR2, TLR4), ACE2, and NRP-1 in the blood samples using quantitative real-time PCR (qRT -PCR).

**Results:** In comparison with the healthy group, all patients exhibited a significant elevation in the serum creatinine, blood urea, cardiac enzymes, and CRP. As well, all studied patients revealed a significant elevation in the expression levels of TLR2, TLR4, ACE2, and NRP-1 mRNA. In all patients, CKMB, ACE2, and NRP-1 mRNA expression levels were positively correlated to both TLR2 and TLR4 expression levels. Moreover, serum creatinine and blood urea were positively correlated to both TLR2 and TLR 4 expression levels in the severe group only.

**Conclusions:** Our study concluded that expression levels for TLR2, TLR4, ACE2, and NRP-1 mRNA in both severe and moderate patients were positively correlated with renal biomarkers and cardiac enzymes. Innate immune markers can be important because they correlate with the severity of illness in COVID-19.

## Introduction

Coronaviruses are described as a large family of enveloped RNA viruses with a single positive strand. They are capable of infecting humans and many animal species. Based on their pathogenicity, human coronaviruses can be categorized into many types.

Highly pathogenic types include severe acute respiratory syndrome coronavirus (SARS-CoV), middle east respiratory syndrome coronavirus (MERS-CoV), and the new severe acute respiratory syndrome coronavirus 2 (SARS-CoV-2) **(Weiss and Navas-Martin, 2005)**. SARS-CoV-2 has led to the severest pandemic of this century with coronavirus disease 2019 (COVID-19). With the world’s second year of the coronavirus pandemic, governments are still struggling to immunize their populations to the herald immunity levels.

On November 5, 2021, the total confirmed cases reached about 248,467,363 with 5,027,128 deaths due to COVID-19 worldwide **(WHO, 2021)**. Originally recognized as respiratory disease, COVID-19 interacts with the cardiovascular system and causes damage to the heart muscle, and results in cardiac and endothelial dysfunction principally via the angiotensin-converting enzyme 2 (ACE2) receptor **(Wang** *et al*., **2020)**. The different tissue tropism between SARS-CoV and SARS-CoV-2 raised the possibility of additional host factors being involved. SARS-CoV-2 advanced protein comprises a cleavage site for protease furin which is not found in SARS-CoV **(Peacock *et al*., 2021)**.

Cantuti-Castelvetri et al. illustrated that neuropilin-1 (NRP-1), which is known to bind furin-cleaved substrates, increases the infectivity of SARS-CoV-2 **(Cantuti** *et al*., **2020**). Viral invasion actuates the host immune system inducing the creation of a large amount of cytokine and interferons to rule out pathogens. Taking the exception of viral DNA/RNA, the viral proteins are also considered as targets of the model recognition receptor. Membrane-linked receptors like (TLRs 1, 2, 4, 6, and 10) refer to viral protein recognition **(Boozari** *et al*., **2019**). Distinct Toll-like receivers (TLRs) play a protective and harmful role for a specific virus. The real mechanistic vision of the pathogenicity of SARS-CoV-2 remains uncertain. This is attributable to the lack of knowledge about why the virus chose humans as its primary host and in what way the virus can escape the innate immune system in humans. In particular, the interactions between human TLRs and viral antigens, and the mechanism of cytokine storms that affect several human organs, are not mostly known. Though, the pathophysiology of COVID-19 usually includes the invasion of the virus into the pulmonary alveoli, usually via the respiratory tract, primarily by way of respiratory droplets, through the airways **(Boozari** *et al*., **2019**). The spike protein, a viral glycoprotein on its capsid, binds to the ACE2 receptor then the RNA genome comes into the host cell through the receptor **(Rothan and Byrareddy 2020)**. After being inside the host cell, viral RNA replicates are formed from mRNA, resulting in quick reproduction of viral RNA and other required structural proteins. Then again, the viral antigens interact with the host’s immune cells and this interaction initiates pro-inflammatory reactions like vasodilation, increased capillary permeability, and gathering of humoral factors **(V’kovski** *et al*., **2021**). All these factors cause the impedance of gas exchange and dyspnea. The exact cause of SARS-CoV-2 and the character of each constituent of the innate and adaptive immune systems are still unidentified (**Sette and Crotty 2021)**. The deficiency of a comprehensive consideration of the pathogenic and immunologic characteristics of the virus made the situation even more alarming to humankind **(V’kovski** *et al*., **2021)**. So, this study aims to evaluate the correlations between mRNA expression levels of Toll-Like Receptors, ACE2 and Neuropilin-1 receptors, and cardiac and renal dysfunction biomarkers in COVID-19.

## Patients and Methods

### Study population

We received laboratory-confirmed medical records and edited data of COVID-19 inpatients stated to the National Health Commission in the period from December 2019 to January 2020. The data deadline for the study was March 2021. All the participants were isolated at Misr International Hospital, Cairo, Egypt, in the period from March 2021 to April 2021. Severe COVID-19 patients are defined as those having hypoxemia (≤ 93% percutaneous oxygen saturation (SpO2) or ≥ 30/min respiratory rate on room air who are on the high-flow nasal cannula or noninvasive mechanical ventilation). Patients not fulfilling the above criteria were weighed as moderate. Informed printed consent was taken from all participants after the ethical committee of the institutional review board was permitted this research.

Cases of Covid19 were confirmed by the presence of positive results for high-throughput sequencing or real-time reverse transcriptase-polymerase chain reaction (RT-PCR) testing of swab samples in the nose and throat **(Raoult** *et al*., **2020)**.

#### Patients

Only cases identified in the laboratory were involved in the study. Eligible COVID-19 patients aged 20-70 years were divided into 2 groups (50 patients each) (**Chen** *et al*., **2020)**.

**Group1;** included patients with moderate symptoms infected by a coronavirus. **Group2;** included patients with severe symptoms infected by a coronavirus and admitted to the intensive care unit (ICU).

### Inclusion and Exclusion Criteria

Healthy individuals were chosen as COVID-19 free volunteers. All patients were recruited between 20-70-year-old and were diagnosed as having COVID-19 according to their PCR and chest CT, as identified by WHO 2020. Key exclusion criteria included hypertensive patients treated with ACE2 inhibitors, pre-existing respiratory disorder, kidney or liver failure, thyroid dysfunction, autoimmune disorders, cerebrovascular diseases, heart diseases, pregnant and lactating women. In addition, patients who receive immunomodulatory drugs or those who have medical conditions such as other infections, malignant tumors, and alcohol abuse.

### The demographic data

Anthropometric variables such as gender and body mass index (BMI) were obtained.

### Blood samples

Serum samples were rapidly separated, aliquoted, and stored at - 40°C until the biochemical measurements. Serum CK-MB and Troponin I were determined by a standard sandwich enzyme-linked immune-sorbent assay (ELISA) Kit gotten from R&D Systems (USA) as guided by the manufacturer. Gene expression of ACE2 mRNA, NRP-1 mRNA, TLR2, TLR4 were spotted in the healthy control, moderate and severe groups in blood samples (**Chen** *et al*., **2020)** using quantitative real-time PCR (qRT-PCR). Gene sequence (5’-3’) procedures were performed conferring to the kit instructions provided in the laboratory assay.

### Laboratory assays

Serum creatinine levels were determined using reagent kits bought from Diamond Diagnostics (Egypt) conferring to the technique of **Henry (1974) (Henry *et al***., **1974)**. Urea concentration was determined according to the technique of **Kaplan (1984) (Kaplan 1984)** using a reagent kit purchased from Diamond Diagnostics (Egypt). Serum lactate dehydrogenase (LDH) activity was measured according to the method of **Buhland Jackson (1978) (Buhland and Jackson 1978)**.

#### RNA isolation and qRT-PCR

Blood samples of all groups were used separately to extract total RNA via TRIzol Reagent purchased from Fermentas, Germany. Then cDNA synthesis was performed utilizing the High-Capacity cDNA Reverse Transcription Kit purchased from Invitrogen, Germany according to the manufacturer’s directions.

Real-time PCR was carried out in a 20 μL system having 10 μL of 1x Sso-Fast Eva Green Supermix (Bio-Rad, Hercules, CA, USA), 2 μL of cDNA, 6 μL of RNase/DNase-free water, and 500nM of the primer pair sequences: NPR1, F:5-AACAACGGCTCGGA CTGGAAGA-3 and R:5-GGTAGATCCTGATGAATCGCGTG -3 (NM001024628); ACE2, F: 5-TCCATTGGTCTTCTGTCACCCG-3 and R: 5-AGACCATCCACCTCCACTTCTC-3 (NM021804.3) β–actin, F: 5-GGAACGGTGAAGGTGACAGCAG-3 and R-5-TGTGGACTTGGGAGAGGACTGG-3 (XM004268956.3); TLR2, F: 5-ATCCTCC AATCAGGCTTCTCT3 and R-5ACACCTCTG TAGGTCACT GTTG3 (NM00131878 9.2); TLR4, F:5-ATATTGACAGGAAACCCCATCCA-3, and R: AGAGAGATTGAGT AGGGGCATTT-3 (NM138554.5); β– actin, F-5AGGAACGGTGAAGGTGACAGCA G-3 and R-5TGTGGACTTGGGAGAGGACTGG-3 (XM00426 8956.3).

The amplification data were analyzed by using the manufacturer’s programmer according to **Livak and Schmittgen** methods **(Livak and Schmittgen 2001)** and the variables were normalized to β–actin.

## Statistical analysis

The result values were accessible as mean ± standard error. SPSS version 20 for Windows (IBM Corp., 2011) was used for data analysis. The one-way analysis of variance (ANOVA) was used to show the statistical differences among groups. This was followed by Duncan’s method for post hoc analysis. By means of the Pearson correlation coefficients method, correlation analysis was estimated between different studied parameters. P-value < 0.05 has been considered as being statistically significant.

## Results

Table 1 shows that the age was significantly higher (p=0.000) in moderate and severe cases compared to healthy controls. Male patients in severe cases were more than female patients. On the other hand, BMI was insignificant (p>0.05) in both infected groups compared to healthy control.

**Table 1:** Demographic and Laboratory findings of severe and moderate COVID-19. Table 1: Data are presented as mean+/-SE. BMI: Body mass index, CRP: C-reactive protein, CKMB: creatinine kinase Myocardial Band, LDH: Lactate dehydrogenase was significantly different at ^**^ *P*< 0.01 level, ^***^ *P*< 0.001 level versus healthy controls, and +*P*< 0.05, +++*P* < 0.001 versus severe group.

|  | Healthy controls | Moderate patients | Severe patients |
| --- | --- | --- | --- |
|  | (n=50) | (n=50) | (n=50) |
| Age (Year) | 34.12±1.24 | 58.24±1.24<sup>***</sup> | 57.36±1.43<sup>***</sup> |
| Gender, no. (%) |  |  |  |
| Male | 25(50%) | 25(50%) | 29 (58%) |
| Female | 25(50%) | 25(50%) | 21 (42%) |
| BMI (kg/m<sup>2</sup>) | 24.25±0.35 | 24.73±0.48 | 25.03±0.52 |
| CRP(mg/dl) | 1.97 ±0.19 | 61.90 ±6.05<sup>+ ***</sup> | 73.92 ±3.42<sup>***</sup> |
| Creatinine (mg/dl) | 0.89 ±0.03 | 0.85 ±0.06<sup>+++</sup> | 1.75 ±0.30<sup>**</sup> |
| Urea (mg/dl) | 25.1 ±0.93 | 48.68 ± 5.49<sup>+++ **</sup> | 79.52 ± 9.14<sup>***</sup> |
| CKMB (ng/ml) | 1.29 ±0.05 | 4.07 ±0.29<sup>***</sup> | 3.55 ±0.20<sup>***</sup> |
| LDH (mg/dl) | 261.6±3.00 | 339.24±25.96<sup>++ **</sup> | 418.7±26.12<sup>***</sup> |
| Troponin I (ng/ml) | 0.02 ±0.002 | 0.03 ±0.003 | 0.04 ±0.01<sup>***</sup> |

Table 1 shows the levels of the kidney function biomarkers and the cardiac enzymes in all groups. Serum creatinine was significantly higher in severe cases (p=0.000) than in moderate and control cases. No significant difference was found between moderate and control cases. The levels of serum high-sensitivity C-reactive protein (hsCRP) (p=0.000), creatinine kinase MB (CKMB) (p=0.000), lactate dehydrogenase (LDH) (p=0.000), and troponin I (p<0.01) were significantly increased among the severe group and moderate group compared to healthy control. Serum (hsCRP) (p=0.000) and levels of lactate dehydrogenase (LDH) (p=0.000) were significantly higher among the severe group compared to the moderate group.

## Discussion

The COVID-19 is a global pandemic. From its starting to spread in December 2019, this viral infection continued to spread internationally. So, efforts are instantly done to stop this virus invasion and guard the world.

To progress an effective treatment and vaccine for COVID-19 infection, decoding the precise immune response to this infection is an essential step. In this case, recovery and disease severity were linked to appropriate immune responses and impaired immune reactions, respectively. Our present data has revealed an increase in serum creatinine and blood urea nitrogen (BUN) compared to patients with normal kidney function. In agreement with our results, **Cheng *et al***. their clinical evidence has shown an elevation in both serum creatinine and BUN in COVID-19 patients, and near 5% of their prospective study in China, patients were diagnosed with acute kidney injury during hospitalization **(Cheng** *et al*., **2020)**.

**Ahmadian *et al***.; **Buonaguro *et al***.; **Huang *et al*. and Wan *et al***. reported that kidney dysfunction mainly acute kidney injury, occurs in cases with COVID-19 within 3 weeks after the beginning of the symptoms; nevertheless, kidney complications are linked to the higher death rate. They concluded that COVID-19 induced organ impairment is mostly facilitated by cytokine storms and that strategies to diminish or eradicate inflammatory cytokines would be effective in avoiding cytokine-induced organ injury. Aiming to the creation of IL1, IL6, TNF, and INF-γ related-inhibitors are reported **(Ahmadian** *et al*., **2021**; **Buonaguro** *et al*., **2020**; **Huang** *et al*., **2020** and **Wan** *et al*., **2020)**. Kidney injury induced by the SARS-CoV-2 virus is probably related to several factors. The virus can infect renal podocytes and cells of the proximal tubules. Disruptive glomerulopathy, leakage of protein in Bowman’s capsule, acute tubular necrosis, and mitochondrial damage based on the angiotensin-converting enzyme 2 (ACE2) pathway **(Fan** *et al*., **2021)**.

Acute kidney injury can result from dysregulation of the immune response, such as cytokine storms and macrophage activation syndrome with lymphopenia **(Ahmadin** *et al*., **2021)**. The development of cytokine storms after viral infection, directly and indirectly, affects the kidneys and can induce sepsis, shock, hypoxia, and rhabdomyolysis. Among the other probable causes of kidney injury induced by COVID-19 is the organ interactions between the pulmonary, cardiac, and renal tissues **(Qian** *et al*., **2021)**.

The present study detected a highly significant upsurge of ACE2 mRNA receptor expressions, NRP-1 mRNA co-receptors, in COVID-19patients.

This finding was consistent with the results of **Imig and Ryan; Cantuti-Castelvetri *et al*.; Daly *et al*.; Freeman and Swartzand** who reported that one of the major receptors for SARS-CoV-2 is ACE2 which is broadly dispersed in the lung, intestinal, liver, heart, vascular endothelium, testis, and kidney cells.

The fact that ACE2 is expressed in respiratory and olfactory epithelial cells at very low protein levels increases the likelihood that other factors are needed to promote virus-host cell communications in the cells that express low ACE2. NRP-1 may be considered as an ACE2 enhancer by facilitating the interface of the virus and ACE2. In addition, TLRs are associated with acute kidney injury, which correlates with the severity of renal disease and inflammatory markers **(Imig and Ryan 2013**; **Cantuti-Castelvetri** *et al*., **2020**; **Daly** *et al*., **2020** and **Freeman and Swartz 2020)**.

Moreover, **Rivero *et al*. and Choudhury and Mukherjee**, concluded these TLRs (2 and 4) can encourage the expression of chemokine in epithelial cells of renal tubules **(Rivero** *et al*., **2009** **Choudhury and Mukherjee 2020)**. Viruses interrelate with precise receptors to enter target cells. SARS-CoV-2 interacts with the angiotensin-converting enzyme 2 receptors (ACE2), which is broadly distributed in the lung, intestinal, liver, heart, vascular endothelium, testis, and kidney cells **(Gheblawi** *et al*., **2020)**. With the help of the type II transmembrane serine protease (TMPRSS2), SARS-CoV-2 comes into the host cells by endocytosis **(Hoffman** *et al*., **2020)**. Zhang et al. revealed that severe COVID-19 patients must show a hyper inflammation nature in comparison to non-severe patients **(Zhang** *et al*., **2020)**.

S1 subunit of the S spike of SARS-CoV-2 has a c-terminal domain that guarantees a very high attraction for the ACE2 receptor **(Huang** *et al*., **2020)**. This declines the ACE2 expression on the cell surface and upsurges inflammation leading to tissues destruction **(Li** *et al*., **2020)**. ACE2 plays an anti-inflammatory role by converting angiotensin II into angiotensin (1-7) **(Issa** *et al*., **(2021)** and reducing vaso-permeability, edema, and pulmonary neutrophil infiltration.

**Ziegler *et al***. recommended that SARS-CoV-2 may raise ACE2 expression and further increase infection **(Ziegler** *et al*., **2020)**. The spike protein of SARS-CoV-2 comprises a cleavage site for the protease furin which is lacking in SARS-CoV. **Cantuti Castelvetri *et al. p***resented that neuropilin-1 (NRP-1), which is identified to bind to furin-cleaving substrates, enhances SARS-CoV-2 infectivity (**Cantuti-Castelvetri** *et al*., **2020)**.

**Daly *et al***. reported that the furin-cleaving S1 fragment of the spike protein directly combines to cell surface NRP-1 and the blocking of this contact with small molecule inhibitors or monoclonal antibodies, will reduce viral invasion in cell culture **(Daly** *et al*., **2020)**. Viral invasion initiates inflammation, stimulation of specialized antigen-presenting cells (APCs) that make the viral peptides presentable to T cells (CD4 and CD8) and stimulates B cells directly. Inflammation is dependent on the first line of innate immune response **(Shah** *et al*., **2020)**. As soon as SARS-CoV-2 enters the host cell, it triggers the pyrin domain containing 3 (NLRP3) inflammasome, one of the Nod-like receptor family. Once the viral RNA interacts with TLRs 3, 7, 8, and 9, A downstream of the NFLκB pathway becomes activated, which augments the production of pro-inflammatory cytokines **(Browne 2020)**. TLR2 and TLR4 are crucial due to their exciting capability to identify different molecular forms of attacking pathogens **(Mukherjee** *et al*., **2016)**. Subsequently, the inflammation twitches and the activated immune cells explode the release of a large number of cytokines. Collectively, these mechanisms participate in augmenting the inflammatory response that plays a major role in the pathogenesis of COVID-19 **(Ye** *et al*., **2020)**. The “cytokine storm syndrome” is believed to be a major underlying factor in the immune-pathogenicity of COVID-19. This storm is responsible for the start of tissue damage, hyper inflammation, and even mortality **(Tang** *et al*., **2020)**. It was activated by the hyperactivation of immune cells leading to a boom of cytokines release. The overexpression of induced IL-10, macrophage IL-1 α, hepatocyte growth factor, IFN-L, IL-3, and were highly related to the severity of the disease **(Tang** *et al*., **2020)**. **Freeman and Swartz** concluded that SARS-CoV-2 is typified by strong and fast stimulation of the innate immune defense, involving triggering of the NLRP3 pathway, and the release of the IL-6 and IL-1 **(Daly** *et al*., **2020**). **Blanco-Melo *et al***. verified that SARS-CoV-2 infection of the epithelial cells of human bronchi gave rise to the expression of numerous cytokines and chemokines like TNF-α, IL-1β, and IL-6 **(Blanco-Melo** *et al*., **2020)**. In addition, our present data showed an elevated troponin I, lactate dehydrogenase (LDH), and CKMB in moderate and severe COVID-19 patients when compared with the control group. COVID-19 has been shown to interrelate and disturb the cardiovascular system, primarily via the ACE2 receptor, resulting in myocardial damage, cardiac damage, and endothelial dysfunction **(Basu-Ray** *et al*., **2020)**. The interface of SARS-CoV-2 with ACE2 can give rise to changes in the ACE2 pathways, causing acute cardiac injury. A few studies propose that SARS-CoV-2 can result in viral myocarditis by direct infection of the myocardium. In many cases, myocardial damage seems to be triggered by increased cardiac metabolic demand which accompanies systemic infections and persistent severe pneumonia-induced hypoxia **(Basu-Ray** *et al*., **2020** **and** **Bugert** *et al*., **2021)**. Moreover, the release of high levels of interleukin 2, 10, 6, 8, and TNF-α can harm several tissues, like vascular endothelium and cardiac myocytes **(Tang** *et al*., **2020)**. Cytokine storms may be associated with the dangerousness of the disease **(Hojyo** *et al*., **2020)**. In a series of situations that affect the kidneys as well as the heart, dysfunction of one organ can encourage dysfunction of the other organ. Chronic or acute systemic conditions can impair the function of these two organs **(Damman and Testani 2015)**. COVID-19-induced acute kidney injury may be related to the crosstalk between the cardiovascular system and the kidney. COVID-19 set off myocarditis to weaken the cardiac output and affect end-organ perfusion. Additionally, the associated proper ventricular disorder produces diastolic disorder and venous congestion that transmit back to the kidney and in addition compromise its blood supply via way of means of growing kidney congestion **(Legrand** *et al*., **2021)**. In addition, acute viral myocarditis together with cytokine cardiomyopathy can induce hypotension, renal venous congestion, and decreased renal blood flow, which can decrease the glomerular filtration rate **(Faour** *et al*., **2022)**.

## Conclusions

Our study concluded that impaired renal biomarkers and cardiac enzymes may be attributed to increased expression levels of TLR2, TLR4, ACE2, and NRP-1 mRNA in both severe and moderate COVID-19 patients. So. it is critical to recognize the molecular mechanisms and focus on the vital molecules involved in the pathogenicity of the diseases to design novel drugs to control and prevent the disease. By blocking the virus access pathways including the viral receptors and regulating immune responses we can reduce multi-organ dysfunction induced by COVID-19.

## Data Availability

All data produced in the present study are available upon reasonable request to the author

## Supporting information

**Fig 1:**
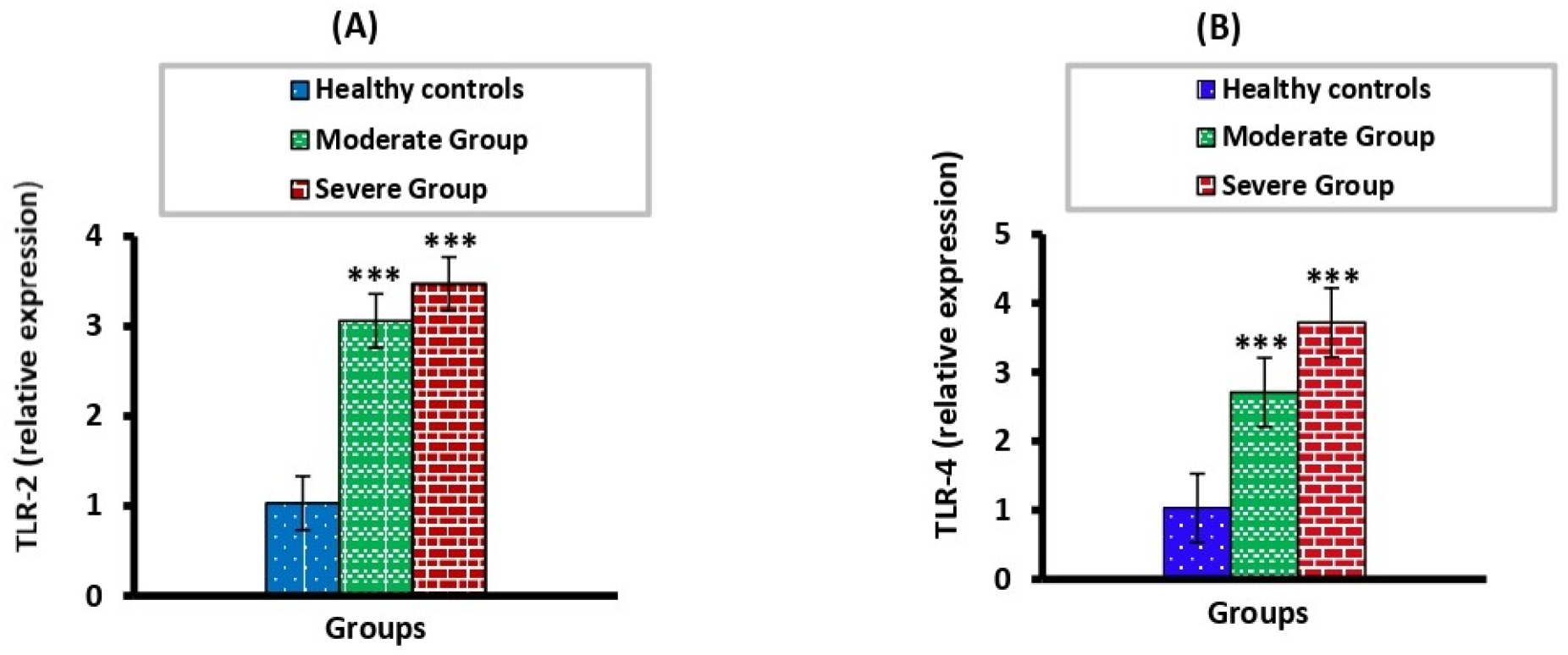
**(A, B)** shows a comparison between the relative expression of TLR-2 and TLR-4 in all studied groups. There were significant (*p*=0.000), (*p*=0.000) increases respectively, in TLR2, TLR4 expression levels in moderate and severe cases compared to healthy controls. **Fig 1: (A)** TLR2; Toll-like receptor 2 and **(B)** TLR4; Toll-like receptor 2 in moderate and severe groups compared to healthy controls.

**Fig 2:**
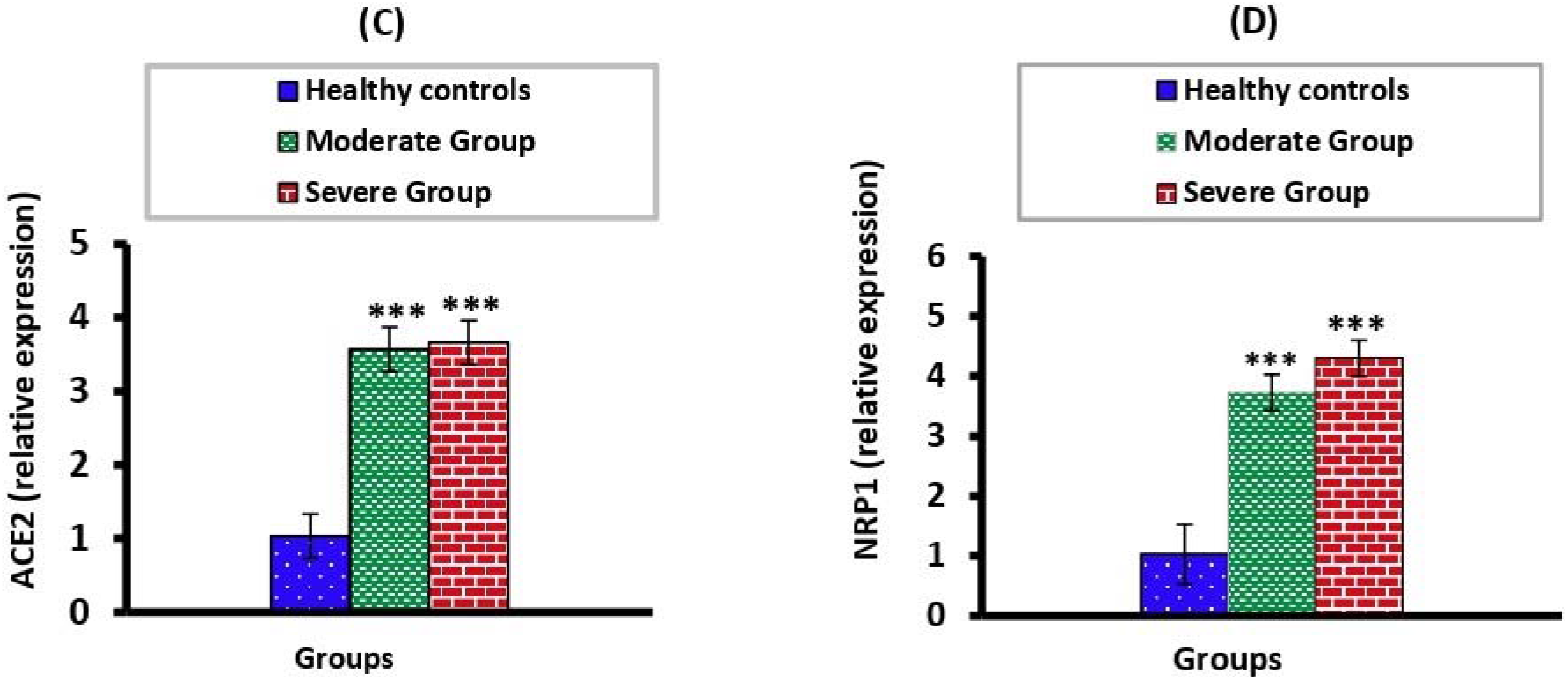
**(C, D)** shows a comparison between the relative ACE2 and NRP-1 mRNA expression levels. This figure exhibited that ACE2 and NRP-1 mRNA expression levels were significantly (p=0.000, p=0.000 respectively), increased in the moderate group and severe group compared to the healthy controls group. **Fig 2: (C)** ACE2; Angiotensin-converting enzyme-2 and **(D)** NPR-1; Neuropilin-1 in moderate and severe groups compared to healthy control

**Fig 3:**
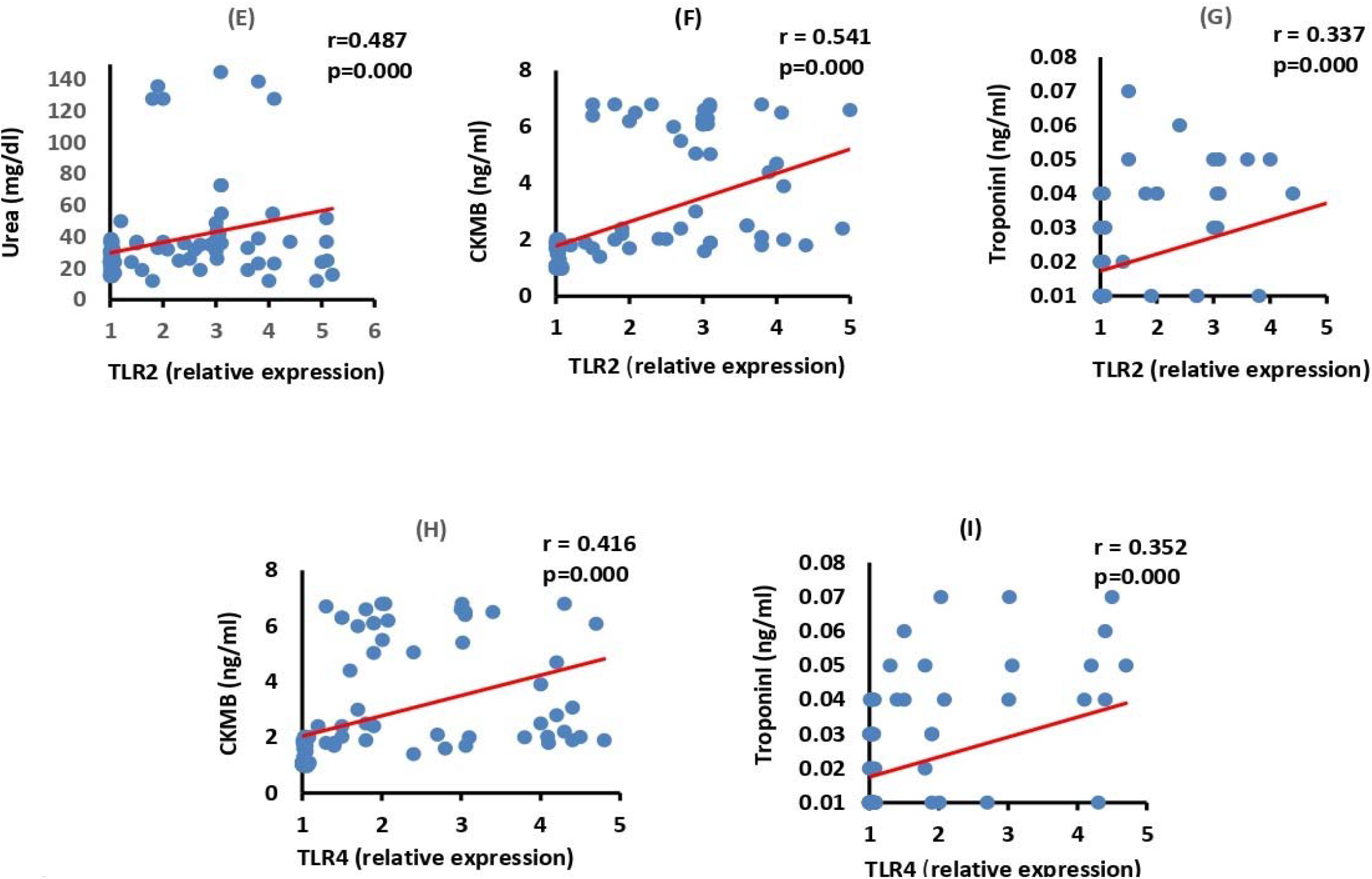
**(E-I) in the moderate group**, shows a significant positive correlation between urea and TLR2 (*p*=0.000). Also, positively correlated between CKMB and Troponin I with both TLR2 (p*=*0.000), (p*<*0.05) and TLR4 (p*=*0.000, p*=*0.000, respectively). **Fig 3: (E-I)** Correlations among the moderate group, between TLR2 with (E) urea, (F) CKMB, (G) Troponin I and TLR4 with (H) CKMB, (I) Troponin I. Correlation was significant ** at the 0.01 level, *** at the 0.001 level. CKMB: creatinine kinase myocardial band; TLR2, 4: Toll-like receptor 2, 4.

**Fig 4:**
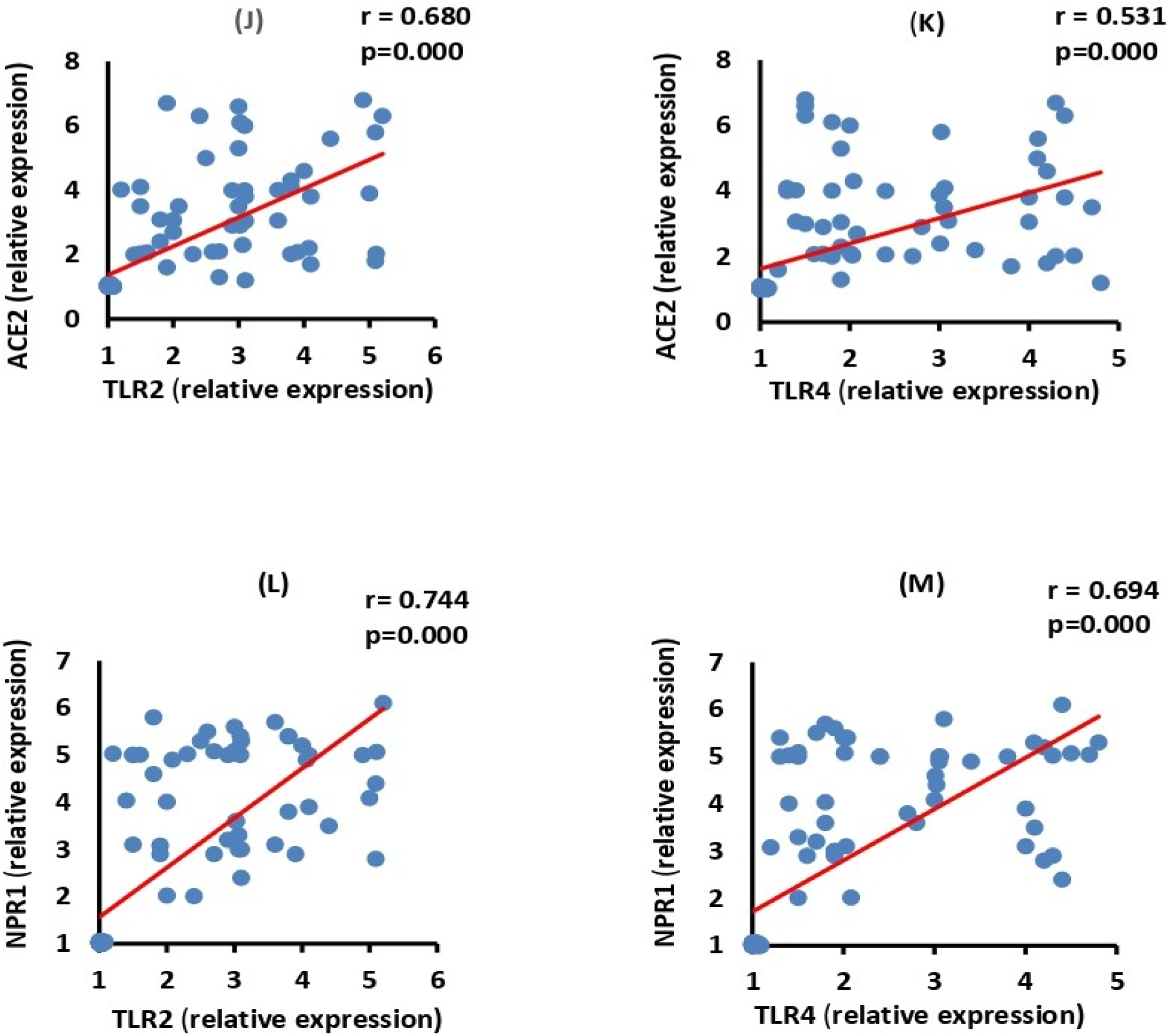
**(J-M)** ACE2 and NRP-1 mRNA expression levels, exhibited a significant positive (p*=*0.000) correlation with TLR2, TLR4 in the moderate group. **Fig 4: (J-M)** Correlations among the moderate group, between ACE2 with (J) TLR2, (K) TLR4, and NRP-1 with (L) TLR2, (M) TLR4. The correlation was significant ** at the 0.01 level, *** at the 0.001 level. ACE2: Angiotensin-converting enzyme; NPR1: Neuroplini-1 and TLR2, 4: Toll-like receptor 2, 4.

**Fig 5:**
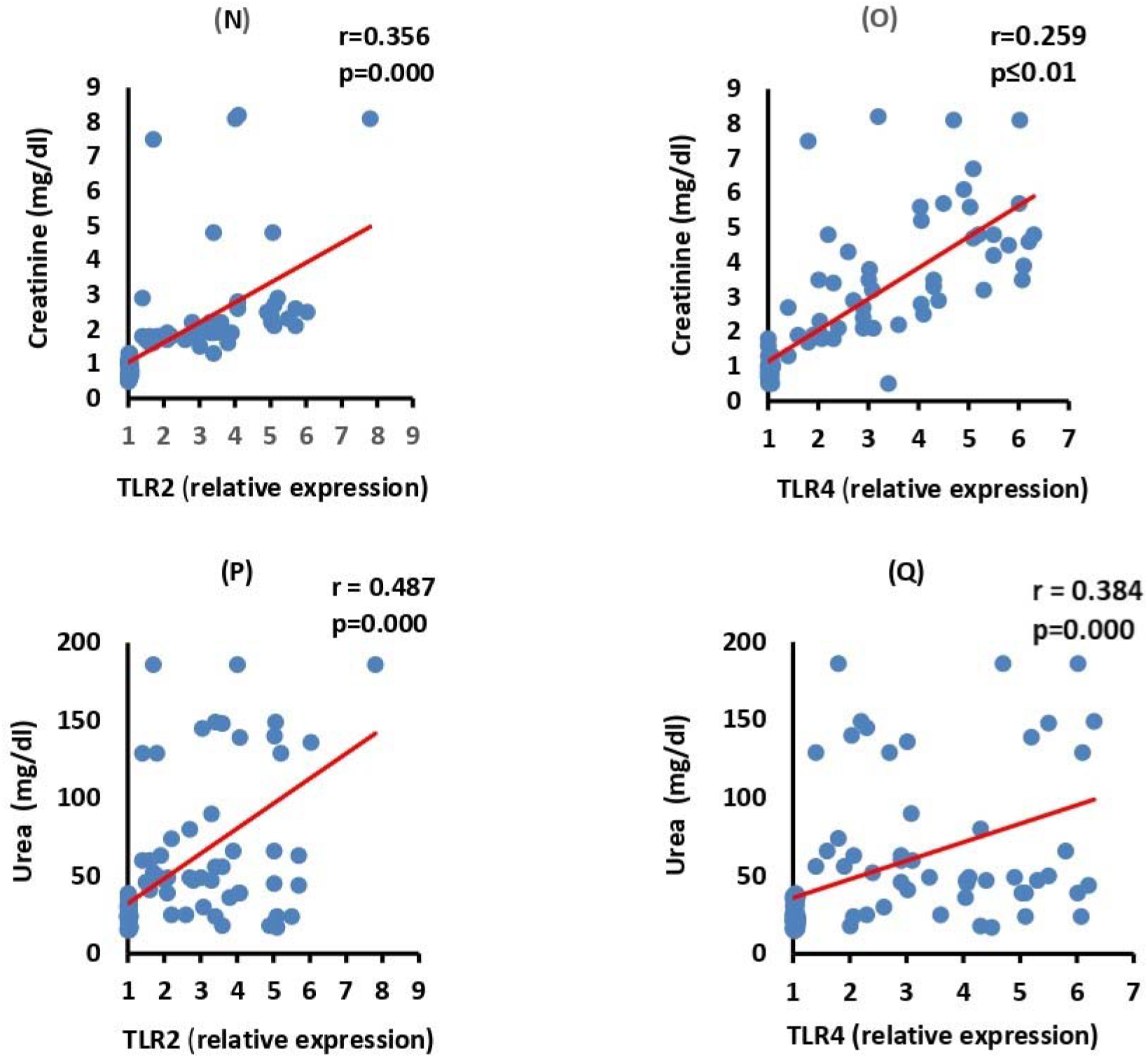
**(N-Q) in the severe group**, both serum creatinine and urea were positively correlated to TLR2 and TLR4 (p*<*0.05). **Fig 5: (N-Q)** Correlations among the severe group, between creatinine with (N) TLR2, (O) TLR4, and urea with (P) TLR2, (Q) TLR4. The correlation was significant ** at the 0.01 level, *** at the 0.001 level. TLR2, 4: Toll-like receptor 2, 4.

**Fig 6:**
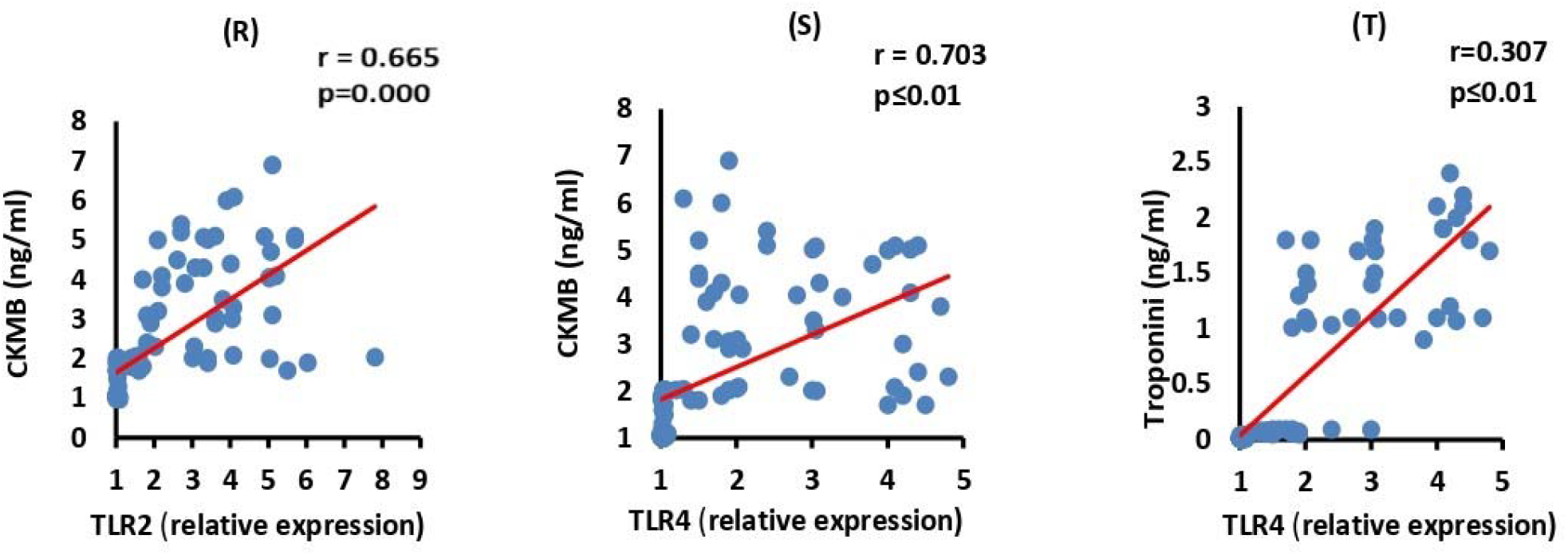
**(R-T)** CKMB showed a positive correlation with TLR2 (p*=*0.000) and TLR4 (p<0.05). Furthermore, Troponin I observed a positive correlation with TLR4 (p*<*0.05) among the severe group. **Fig 6: (R-T)** Correlations among the severe group, between CKMB with (R) TLR2, (S) TLR4 and Troponin I with (T) TLR4. The correlation was significant ** at the 0.01 level, *** at the 0.001 level. CKMB: creatinine kinase myocardial band; TLR2, 4: Toll-like receptor 2, 4.

**Fig 7:**
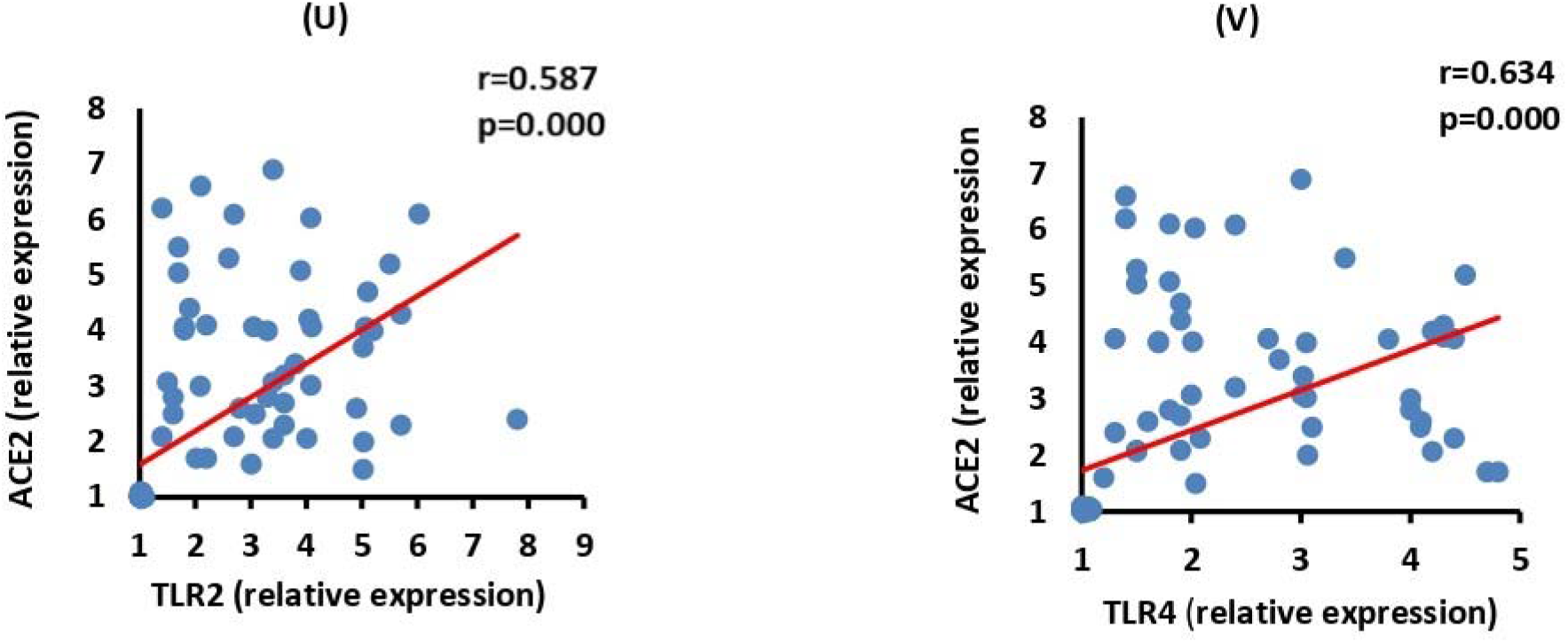
**(U, V)** shows that ACE2 mRNA expressions were positively (p=0.000) correlated with TLR2 and TLR4 in the severe group. **Fig 7: (U, V)** Correlations among the severe group, between ACE2 with (U) TLR2, (V) TLR4. The correlation was significant ** at the 0.01 level, *** at the 0.001 level. ACE2: Angiotensin-converting enzyme; TLR2, 4: Toll-like receptor 2, 4.

**Fig 8:**
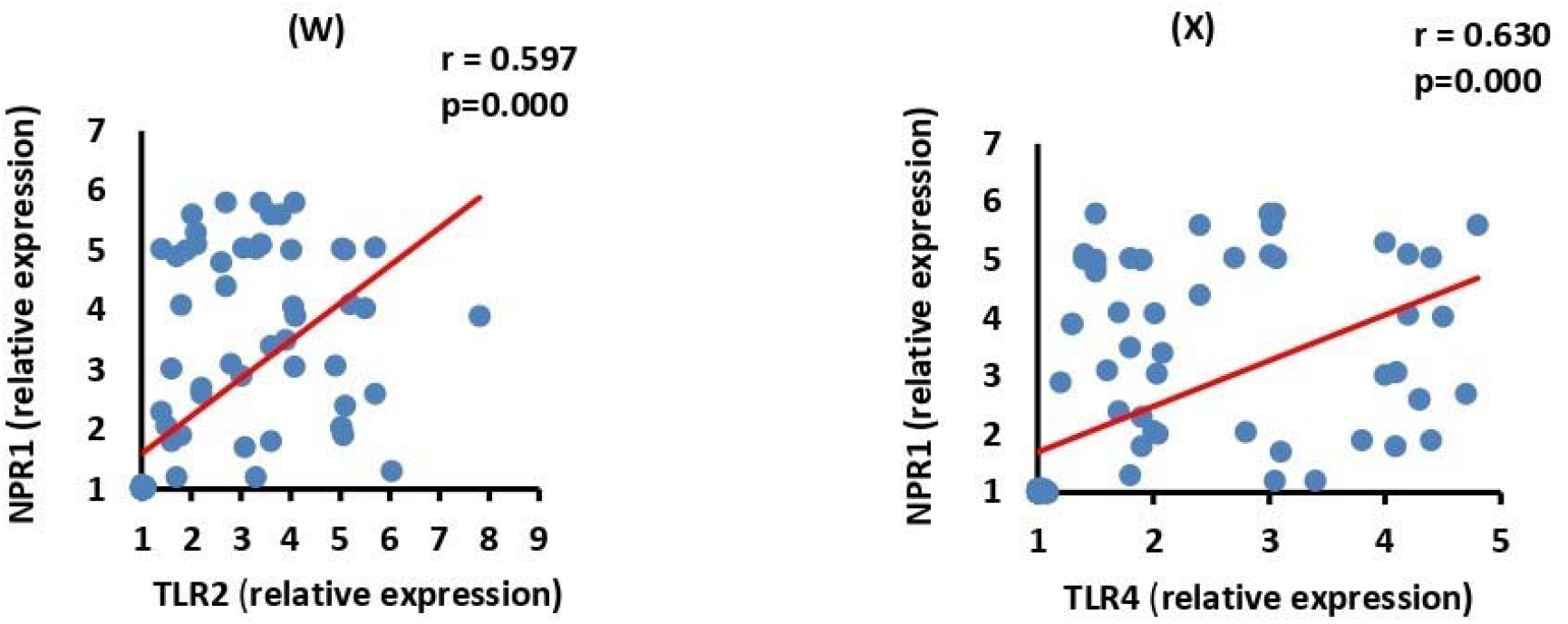
**(W, X)** shows that NRP-1 mRNA expressions were positively (p=0.000) correlated to TLR2 and TLR4 in the severe group. **Fig 8: (W, X)** Correlations among the severe group, between NRP-1 with (W) TLR2, (X) TLR4. The correlation was significant ** at the 0.01 level, *** at the 0.001 level. ACE2: Angiotensin-converting enzyme; NPR-1: Neuroplini-1; TLR2, 4: Toll-like receptor 2, 4.

## Acknowledgments

The authors would like to thank Taif University, Taif, Saudi Arabia for supporting this research (Taif University researchers supporting project number: TURSP-2020/127). Thank you very much Dr. Ahmed Abdelhafeiz consultant of Medical Biochemistry - Al Azhar University.

## Funding

“This work was supported by [Taif University, Taif, Saudi Arabia] (Grant number [TURSP-2020/127]. The author, Basem Hassan Elesawy has received research support from Taif University, Taif, Saudi Arabia.”

### Ethics Statement

The studies were reviewed and approved by the Ethics Committee of Faculty of Post-graduate studies for Advanced Science, Beni-Suef University, Egypt, and approval from Misr International Hospital was obtained.

### Consent to participate

All patients/participants or their relatives provided their written informed consent to participate in this study.

### Authors contribution

Conceptualization: [Maged Abdallah, Osama M. Ahmed]; Methodology: [Rabab Hussain Sultan, Amr E. Ahmed, Basem Hassan Elesawy, Maged Abdallah]; Formal analysis and investigation: [Hebatallah Hany Assal, Tarek Mohamed Ali, Osama M. Ahmed]; Writing - original draft preparation: [Rabab Hussain Sultan, Tarek Mohamed Ali, Osama M. Ahmed]; Writing - review and editing: [Tarek Mohamed Ali, Osama M. Ahmed, Hebatallah Hany Assal]; Funding acquisition: [Basem Hassan Elesawy]; Resources: [Rabab Hussain Sultan, Maged Abdallah, Tarek Mohamed Ali, Hebatallah Hany Assal, Amr E. Ahmed, Basem Hassan Elesawy]; Supervision: [Osama M. Ahmed, Maged Abdallah, Hebatallah Hany Assal, Amr E. Ahmed].

### Consent to publish

All authors agreed with the content and that all gave explicit consent to submit and that they obtained consent from the responsible authorities at the institute/organization where the work has been carried out **before** the work is submitted.

### Data and availability of material

All authors guarantee transparency on the re-use of material and that there is no unpublished material (for example manuscripts in press) included in the manuscript

## References

Ahmadian, E.; Hosseiniyan Khatibi, S. M.; Razi Soofiyani, S.; Abediazar, S.; Shoja, M. M.; Ardalan, M. and Zununi Vahed, S. (2021): COVID-19and kidney injury: Pathophysiology and molecular mechanisms. Reviews in medical virology, 31(3), e2176.

Basu-Ray, I.; Almaddah, N. K.; Adeboye, A. and Soos, M. P. (2021): Cardiac Manifestations Of Coronavirus (COVID-19). In StatPearls. StatPearls Publishing.

Blanco-Melo, D.; Nilsson-Payant, B. E.; Liu, W. C.; Uhl, S.; Hoagland, D.; Møller, R.; Jordan, T. X.; Oishi, K.; Panis, M.; Sachs, D.; Wang, T. T.; Schwartz, R. E.; Lim, J. K.; Albrecht, R. A., and tenOever, B. R. (2020): Imbalanced Host Response to SARS-CoV-2 Drives Development of COVID-19. Cell, 181(5), 1036–1045.e9.

Boozari, M.; Butler, A.E. and Sahebkar A. (2019): Impact of curcumin on toll-like receptors. J Cell Physiol. 2019; 234(8):12471–12482.

Browne E. P. (2020): The Role of Toll-Like Receptors in Retroviral Infection. Microorganisms, 8(11), 1787.

Bugert, C. L.; Kwiat, V.; Valera, I. C.; Bugert, J. J. and Parvatiyar, M. S. (2021): Cardiovascular Injury Due to SARS-CoV-2. Current clinical microbiology reports, 1–11. Advance online publication.

Buhl, S. N. and Jackson, K. Y. (1978): Optimal conditions and comparison of lactate dehydrogenase catalysis of the lactate-to-pyruvate and Pyruvate-to-lactate in human serum at 25, 30-and 37-degree C. Clin. Chem., 24: 828.

Buonaguro, F. M.; Ascierto, P. A.; Morse, G. D.; Buonaguro, L.; Puzanov, I.; Tornesello, M. L.; Bréchot, C. and Gallo, R. C. (2020). Covid-19: Time for a paradigm change. Reviews in medical virology, 30(5), e2134.

Cantuti-Castelvetri, L.; Ojha, R.; Pedro, L. D.; Djannatian, M.; Franz, J.; Kuivanen, S.; van der Meer, F.; Kallio, K.; Kaya, T.; Anastasina, M.; Smura, T.; Levanov, L.; Szirovicza, L.; Tobi, A.; Kallio-Kokko, H.; Österlund, P.; Joensuu, M.; Meunier, F. A.; Butcher, S. J.; Winkler, M. S. and Simons, M. (2020): Neuropilin-1 facilitates SARS-CoV-2 cell entry and infectivity. Science (New York, N.Y.), 370(6518), 856–860.

Chen, G.; Wu, D.; Guo, W.; Cao, Y.; Huang, D.; Wang, H.; Wang, T.; Zhang, X.; Chen, H.; Yu, H.; Zhang, X.; Zhang, M.; Wu, S.; Song, J.; Chen, T.; Han, M.; Li, S.; Luo, X.; Zhao, J. and Ning, Q. (2020): Clinical and immunological features of severe and moderate coronavirus disease 2019. The Journal of clinical investigation, 130(5), 2620–2629.

Cheng, Y.; Luo, R.; Wang, K.; Zhang, M.; Wang, Z.; Dong, L.; Li, J.; Yao, Y.; Ge, S. and Xu, G. (2020): kidney disease is associated with in-hospital death of patients with COVID-19. Kidney International, 97(5), 829–838.

Choudhury, A. and Mukherjee, S. (2020): In silico studies on the comparative characterization of the interactions of SARS-CoV-2 spike glycoprotein with ACE2 receptor homologs and human TLRs. Journal of medical virology, 92(10), 2105–2113.

Daly, J. L.; Simonetti, B.; Klein, K.; Chen, K. E.; Williamson, M. K.; Antón-Plágaro, C.; Shoemark, D. K.; Simón-Gracia, L.; Bauer, M.; Hollandi, R.; Greber, U. F.; Horvath, P.; Sessions, R. B.; Helenius, A.; Hiscox, J. A.; Teesalu, T.; Matthews, D. A.; Davidson, A. D.; Collins, B. M.; Cullen, P. J. and … Yamauchi, Y. (2020): Neuropilin-1 is a host factor for SARS-CoV-2 infection. Science (New York, N.Y.), 370(6518), 861–865.

Damman, K. and Testani, J. M. (2015): The kidney in heart failure: an update. European heart journal, 36(23), 1437–1444.

Fan, C.; Lu, W.; Li, K.; Ding, Y. and Wang, J. (2021): ACE2 Expression in Kidney and Testis May Cause Kidney and Testis Infection in COVID-19Patients. Frontiers in medicine, 7, 563893.

Faour, W. H.; Choaib, A.; Issa, E., Choueiry, F. E.; Shbaklo, K.; Alhajj, M.; Sawaya, R. T.; Harhous, Z.; Alefishat, E. and Nader, M. (2022): Mechanisms of COVID-19-induced kidney injury and current pharmacotherapies. Inflammation research: official journal of the European Histamine Research Society … [et al.], 71(1), 39–56.

Freeman, T. L., and Swartz, T. H. (2020): Targeting the NLRP3 Inflammasome in Severe COVID-19. Frontiers in immunology, 11, 1518.

Gheblawi, M.; Wang, K.; Viveiros, A.; Nguyen, Q.; Zhong, J. C.; Turner, A. J.; Raizada, M. K.; Grant, M. B. and Oudit, G. Y. (2020): Angiotensin-Converting Enzyme 2: SARS-CoV-2 Receptor and Regulator of the Renin-Angiotensin System: Celebrating the 20th Anniversary of the Discovery of ACE2. Circulation Research, 126(10), 1456–1474.

Henry T.J. 2nd ed. Harper and Row Publishers (1974): New York: Clinical Chemistry Principles and techniques.[Google Scholar]

Hoffmann, M.; Kleine-Weber, H.; Schroeder, S.; Krüger, N.; Herrler, T.; Erichsen, S.; Schiergens, T. S.; Herrler, G.; Wu, N. H.; Nitsche, A.; Müller, M. A.; Drosten, C. and Pöhlmann, S. (2020): SARS-CoV-2 Cell Entry Depends on ACE2 and TMPRSS2 and Is Blocked by a Clinically Proven Protease Inhibitor. Cell, 181(2), 271–280.e8.

Hojyo, S.; Uchida, M.; Tanaka, K.; Hasebe, R.; Tanaka, Y.; Murakami, M. and Hirano, T. (2020): How COVID-19induces cytokine storm with high mortality. Inflammation and regeneration, 40, 37.

Huang, C.; Wang, Y.; Li, X.; Ren, L.; Zhao, J.; Hu, Y.; Zhang, L.; Fan, G.; Xu, J.; Gu, X.; Cheng, Z.; Yu, T.; Xia, J.; Wei, Y.; Wu, W.; Xie, X..; Yin, W.; Li, H.; Liu, M.; Xiao, Y. and … Cao, B. (2020): Clinical features of patients infected with 2019 novel coronavirus in Wuhan, China. Lancet (London, England), 395(10223), 497–506.

Huang, Y.; Yang, C.; Xu, X. F.; Xu, W. and Liu, S. W. (2020): Structural and functional properties of SARS-CoV-2 spike protein: potential antivirus drug development for COVID-19. Acta pharmacologica Sinica, 41(9), 1141–1149.

Imig, J. D., and Ryan, M. J. (2013): Immune and inflammatory role in renal disease. Comprehensive Physiology, 3(2), 957–976.

index in coronavirus (COVID-19) infected patients. British journal of haematology, 189(3), 428–437.

Issa, H.; Eid, A. H.; Berry, B.; Takhviji, V.; Khosravi, A.; Mantash, S.; Nehme, R.; Hallal, R.’ Karaki, H.; Dhayni, K.; Faour, W. H.; Kobeissy, F.; Nehme, A. and Zibara, K. (2021): Combination of Angiotensin (1-7) Agonists and Convalescent Plasma as a New Strategy to Overcome Angiotensin-Converting Enzyme 2 (ACE2) Inhibition for the Treatment of COVID-19. Frontiers in medicine, 8, 620990.

Kaplan A. (1984): Urea Clin Chem. Pbl. The C.V. Mosby Co. St Louis. Toronto. Princeton, PP. 1257–1260 and 437 and 418.

Legrand, M.; Bell, S.; Forni, L.; Joannidis, M.; Koyner, J. L.; Liu, K. and Cantaluppi, V. (2021): Pathophysiology of COVID-19-associated acute kidney injury. Nature reviews. Nephrology, 17(11), 751–764.

Li, Y.; Zhou, W.; Yang, L.; You, R. (2020): Physiological and pathological regulation of ACE2, the SARS-CoV-2 receptor. Pharmacological Research, 157, 104833.

Livak, K.J. and Schmittgen, T.D. (2001): Analysis of relative gene expression data using real-time quantitative PCR and the 2(-Delta Delta C (T)) method. Methods. 2001; 25(4): 402–408.

Mukherjee, S.; Karmakar, S. and Babu, S. P. (2016): TLR2 and TLR4 mediated host immune responses in major infectious diseases: a review. The Brazilian journal of infectious diseases: an official publication of the Brazilian Society of Infectious Diseases, 20(2), 193–204.

Peacock, T. P., Goldhill, D. H., Zhou, J., Baillon, L., Frise, R., Swann, O. C., Kugathasan, R., Penn, R., Brown, J. C., Sanchez-David, R. Y., Braga, L., Williamson, M. K., Hassard, J. A., Staller, E., Hanley, B., Osborn, M., Giacca, M., Davidson, A. D., Matthews, D. A., & Barclay, W. S. (2021). The furin cleavage site in the SARS-CoV-2 spike protein is required for transmission in ferrets. Nature microbiology, 6(7), 899–909.

Qian, J. Y.; Wang, B.; Lv, L. L. and Liu, B. C. (2021): Pathogenesis of Acute Kidney Injury in Coronavirus Disease 2019. Frontiers in physiology, 12, 586589. https://doi.org/10.3389/fphys.2021.586589.

Raoult, D.; Zumla, A.; Locatelli, F.; Ippolito, G. and Kroemer, G. (2020): Coronavirus infections: epidemiological, clinical and immunological features and hypotheses. Cell Stress. 2020; 4:66–75. 10.15698/cst2020.04.216

Rivero, A.; Mora, C.; Muros, M.; García, J.; Herrera, H. and Navarro-González, J. F. (2009): Pathogenic perspectives for the role of inflammation in diabetic nephropathy. Clinical science (London, England: 1979), 116 (6), 479–492.

Rothan, H.A., and Byrareddy, S.N. (2020): The epidemiology and pathogenesis of coronavirus disease (COVID-19) outbreak. J Autoimmun. 2020;109:102433 10.1016/j.jaut.2020.102433

Sette, A., and Crotty, S. (2021): Adaptive immunity to SARS-CoV-2 and COVID-19. Cell, 184(4), 861–880.

Shah, V. K.; Firmal innate immune responses, P.; Alam, A.; Ganguly, D. and Chattopadhyay, S. (2020): Overview of Immune Response During SARS-CoV-2 Infection: Lessons From the Past. Frontiers in immunology, 11, 1949.

Tang, Y.; Liu, J.; Zhang, D.; Xu, Z.; Ji, J. and Wen, C. (2020): Cytokine Storm in COVID-19: The Current Evidence and Treatment Strategies. Frontiers in immunology, 11, 1708.

V’kovski, P., Kratzel, A., Steiner, S., Stalder, H., and Thiel, V. (2021): Coronavirus biology and replication: implications for SARS-CoV-2. Nature reviews. Microbiology, 19(3), 155–170.

Wan, S.; Yi, Q.; Fan, S.; Lv, J.; Zhang, X.; Guo, L.; Lang, C.; Xiao, Q.; Xiao, K.; Yi, Z.; Qiang, M.; Xiang, J.; Zhang, B.; Chen, Y. and Gao, C. (2020): Relationships among lymphocyte subsets, cytokines, and the pulmonary inflammation

Wang, D.; Hu, B.; Hu, C.; Zhu, F.; Liu, X.; Zhang, J.; Wang, B.; Xiang, H.; Cheng, Z.; Xiong, Y, Zhao, Y.; Li, Y.; Wang, X. and Peng, Z. (2020): Clinical Characteristics of 138 Hospitalized Patients With 2019 Novel Coronavirus-Infected Pneumonia in Wuhan, China. JAMA. 2020 Mar 17; 323(11):1061–1069.

Weiss, S.R. and Navas-Martin S. (2005): Coronavirus pathogenesis and the emerging pathogen severe acute respiratory syndrome coronavirus. Microbiol Mol Biol Rev. 2005 Dec; 69(4):635–64.

World Health Organization, (2021): WHO Coronavirus (COVID-19) Dashboard.

Ye, Q.; Wang, B. and Mao, J. (2020): The pathogenesis and treatment of the ‘Cytokine Storm’ in COVID-19. The Journal of infection, 80(6), 607–613.

Zhang, J. J.; Dong, X.; Cao, Y. Y.; Yuan, Y. D.; Yang, Y. B.; Yan, Y. Q.; Akdis, C. A. and Gao, Y. D. (2020): Clinical characteristics of 140 patients infected with SARS-CoV-2 in Wuhan, China. Allergy, 75(7), 1730–1741.

Ziegler, C.; Allon, S. J.; Nyquist, S. K.; Mbano, I. M.; Miao, V. N.; Tzouanas, C. N.; Cao, Y.; Yousif, A. S.; Bals, J.; Hauser, B. M.; Feldman, J.; Muus, C.; Wadsworth, M. H.; 2nd, Kazer, S. W.; Hughes, T. K.; Doran, B.; Gatter, G. J.; Vukovic, M.; Taliaferro, F.; Mead, B. E. and … HCA Lung Biological Network (2020): SARS-CoV-2 Receptor ACE2 Is an Interferon-Stimulated Gene in Human Airway Epithelial Cells and Is Detected in Specific Cell Subsets across Tissues. Cell, 181(5), 1016–1035.e19.

